# Which outcomes should be included in a core outcome set for capturing and measuring doctor well-being? A Delphi study

**DOI:** 10.1101/2024.04.11.24305668

**Authors:** Gemma Simons, Naomi Klepacz, David S. Baldwin

**Author notes:** Corresponding author: Dr Naomi Klepacz.

## Abstract

**Objectives:** To develop a core outcome set (COS) to capture and measure the well-being of doctors working in the NHS.

**Design:** An online Delphi study.

**Setting:** United Kingdom National Health Service.

**Participants:** Participants from four stakeholder groups: i) those who might use the COS in research, ii) organisations that measure/capture NHS staff wellbeing, iii) professionals with experience managing NHS staff wellbeing, and iv) NHS doctors, were identified through authorship of relevant publications, attendee lists of doctor well-being conferences and meetings, professional bodies, participation in a previous study and recommendations from others. They were recruited via email.

**Method:** A two-stage process: 1) creating a list of 43 wellbeing outcomes informed by a systematic review of wellbeing measurement instruments, a survey of UK doctors and 2 doctor engagement workshops, and 2) an online modified Delphi study (with two rounds) to reach consensus. Outcomes were rated on a 9-point Likert scale; ‘consensus’ was reached when ≤75% agreed that an outcome was critical for inclusion in the COS.

**Results:** Fifty-two participants completed both Delphi rounds. Seven wellbeing outcomes met the threshold for inclusion in the COS: General wellbeing, Health, Personal safety, Job satisfaction, Morale, Life work balance, and Good clinical practice.

**Conclusion:** Use of the COS has the potential to reduce heterogeneity and standardise the capture and measurement of doctor well-being and ensure outcomes important to all stakeholders are reported.

**Trial registration:** This study was prospectively registered with the COMET initiative www.comet-initiative.org (Registration: 1577)

## STRENGTHS AND LIMITATIONS

- This is the first study to develop a core outcome set (COS) for the capture and measurement of doctor well-being.
- A salutogenic and consensus approach was used to achieve agreement between four stakeholder groups.
- Without an internationally agreed definition of doctor well-being, identification and classification of well-being outcomes were subjective.
- There is potential to use the COS with other groups of healthcare professionals.
- Further research is needed to identify agreed measurement tools.

Doctors are increasingly experiencing high workloads and challenging working conditions and, consequently, are reporting high levels of stress, anxiety, depression,^1^ emotional distress, burnout risk^2^ ^3^ and suicidal feelings. ^4^ ^5^ This negatively impacts patient care quality, safety and satisfaction^6–8^ and leads to declining job satisfaction and doctors leaving the workforce.^9^ The 2022 NHS staff survey^10^ found that 46% of doctors felt unwell in the last 12 months because of work-related stress, and 40% often or always found their work emotionally exhausting. Emphasis is often placed on doctors to be more resilient, with stigma and a fear of potential repercussions preventing doctors from speaking up about their well-being.^11^ However, there is an emerging consensus that some aspects of doctors’ training, working conditions and organisational support negatively impact well-being.^5^ The well-being of doctors significantly impacts workforce planning, cost, healthcare quality and patient outcomes.^12^ Dissatisfaction with role/place of work or NHS culture was cited as the top reason for leaving the workforce in a General Medical Council survey,^13^ with burnout/work-related stress as the third most cited reason behind retirement. Poor mental well-being of staff is estimated to cost the NHS at least £12.1 billion per year; tackling poor mental well-being and reducing the number of staff leaving the NHS could save up to £1 billion.^14^ The UK’s health system prioritises patient care - often over staff well-being - but long-term patient care and safety depend on staff well-being.^15^

The need to address doctor well-being is well recognised, with government and industry reports highlighting the need for improvement (e.g., ^2^ ^8^ ^11^ ^15–18^). While recognising the urgent need to address doctors’ well-being, these reports often fail to operationalise well-being or specify the outcome or measurement tools required to gauge the success of their recommendations. For example, the ‘NHS Long Term Plan’^19^ aims to make the NHS ‘the best place to work’ but provides little detail on implementation or how success will be captured or measured. The NHS long term workforce plan^20^ commits to implementing actions from the NHS People Plan^21^ to ensure that staff have access to well-being services and support; however, the British Medical Association has questioned how this ambition will be made a reality.^22^ Many employers and education deaneries now provide well-being programmes for doctors and implement the NHS health and well-being framework.^23^ However, evidence of the success of these (and similar) programmes – often aimed at individual coping strategies, resilience and productivity – suggests limited effect.^12^ The lack of consensus on what doctor well-being is and how it should be measured means that the monitoring and evaluation of these strategies are inconsistent.

The ongoing and accurate measurement of doctors’ well-being is necessary to understand local and specific needs and ensure the effective delivery of staff services.^17^ It is, therefore, vital for both research and governance to take a consistent data-driven and evidence-based approach to doctors’ well-being, taking account of the many dimensions (i.e., social, cultural, environmental, economic) and levels (i.e., individual, organisational, societal) that comprise this complex issue. However, work has not yet been undertaken to standardise the definition and measurement of doctor well-being. In addition, well-being has often been used interchangeably with, or to describe, mental health, with previous research focusing largely on ‘pathologies’ such as depression, anxiety and burnout rather than positive measures of well-being. Consequently, workplace well-being has become a measure of the absence of mental health disorders. A salutogenic approach^24^ ^25^ that measures positive determinants, context, mechanisms, and individual and group well-being should be preferred when considering doctor well-being.^26^

Our systematic review^27^ found well-being outcomes and measurement tools used in doctor well-being research were heterogeneous, demonstrating the need for a core outcome set (COS). Core outcome sets are consensus minimum groups of outcomes with recommended reliable and valid measurement tools. Reaching agreement among stakeholders – including NHS doctors - ensures a consistent and comprehensive focus, facilitating comparison between organisations through the generation of ‘big-data’, and in doing so, provides decision-makers with the evidence needed to inform future workforce strategies, interventions and actions. We used a salutogenic and consensus-based approach to develop a COS to capture and report on the well-being of doctors in the NHS. To our knowledge, this study represents the first time a non-pathological concept – well-being – has been applied to the Core Outcome Set-Standards for Reporting (COS-STAD) guidance.^28^

## METHODS

### Study Overview

The COS was developed in two stages: 1) the generation of a long list of outcomes, and 2) an online Delphi survey (Figure 1). The study protocol was developed following the Core Outcome Measures in Effectiveness Trials (COMET) criteria^29^ and was prospectively registered with the COMET initiative^30^ (Registration: 1577). The study is reported using the COS-STAD guidance. ^31^

**Figure 1.**
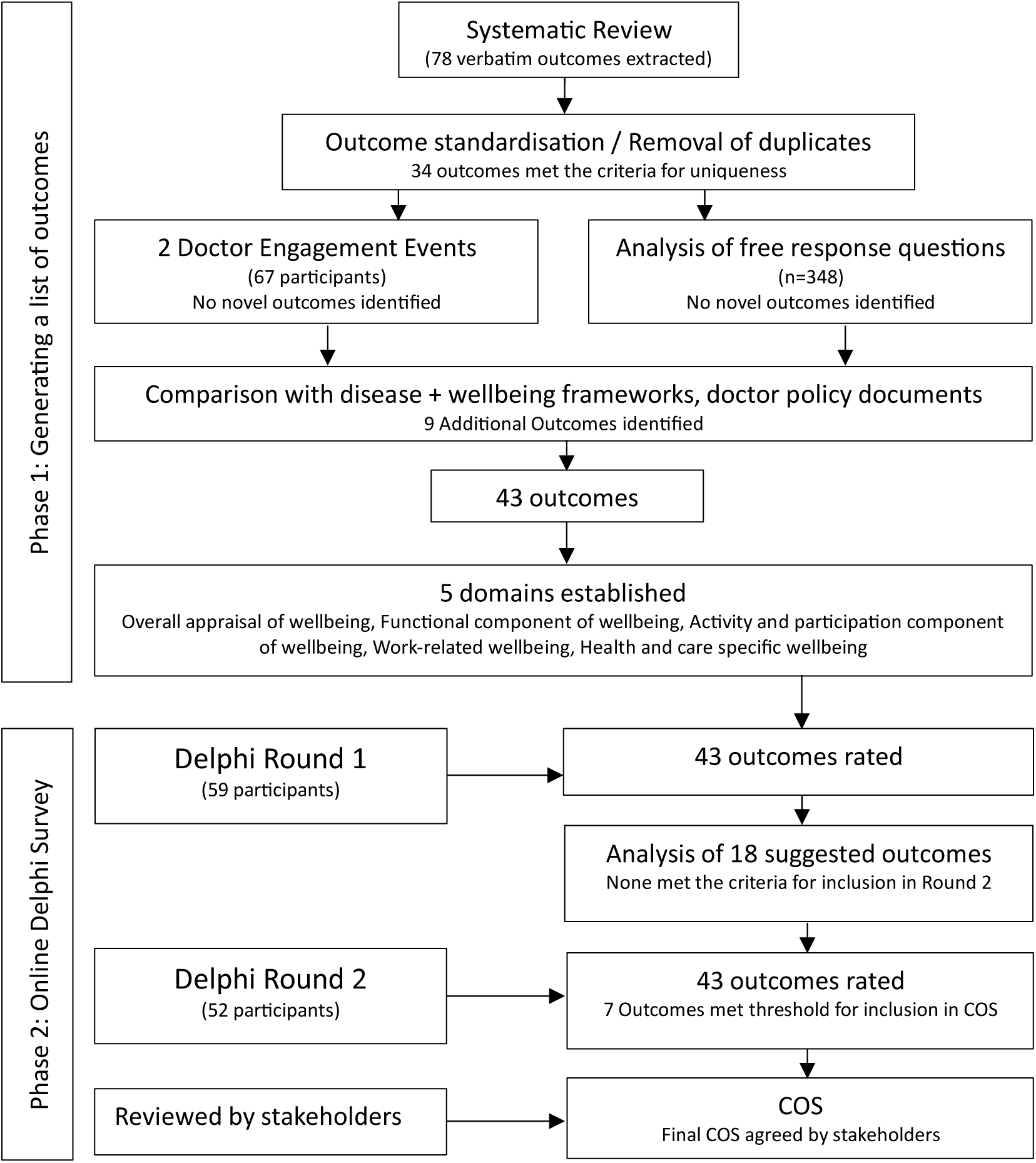
Flow diagram of the process for developing COS.

### Ethical Approval

This study involved human participants and was approved by the University of Southampton Faculty of Medicine Ethics Committee (ERGO 49246). Participants gave informed consent before participating.

### Phase 1: Generating a List of Outcomes

The long list of outcomes was generated from a systematic review of doctor well-being definitions and measures,^27^ local and national doctor involvement/engagement events,^27^ and analysis of free text response questions from a national online survey of doctor well-being.^27^ Inductive constant comparison analysis ^32^ allowed the participants to generate themes. As part of this ‘open coding’ technique, participants’ own words were used for code names, and the themes and meanings were constructed after the data had been collected using convergent thematic analysis.^33^ Finally, any duplicates were removed, and similar concepts were merged. Outcomes that were pathologies, symptoms of pathologies, or negative in nature were removed from the long list. Uniqueness was established when the published definition of an outcome differed conceptually from those of other outcomes.^34^ Outcomes identified as unique were compared to the outcomes and domains in health, health-related quality of life, disease, and well-being frameworks, ^35–38^ and doctor policy documents ^21^ ^39–43^ to identify any further key concepts. Plain English descriptions – presented as ‘help text’ in the Delphi survey – were written for each outcome, guided by the published literature (See supplementary materials 1). We established relevant domains by grouping thematically similar outcomes that captured a broader concept using a well-being framework developed for this study.^27^ ‘Think aloud’ interviews were conducted (n=3 doctors, n=2 psychologists) to review the uniqueness of the outcomes and the clarity, conciseness, and accessibility of the ‘help-text’ definitions for stakeholders.

### Phase 2: Online Delphi Survey

The long list of outcomes was used to populate an online Delphi survey, delivered using the DelphiManager platform.^44^ The Delphi approach is a method widely used in developing core outcome sets.^44^ ^45^ The method aims to achieve consensus through the collection and synthesis of stakeholder opinions.

#### Stakeholder recruitment

A purposive sampling method was used to recruit participants from four stakeholder groups: i) those who could use the core outcome set in research, ii) representatives of organisations that measure doctor well-being in the NHS, iii) professionals with experience in managing doctor well-being, and iv) doctors working in the NHS (all specialities and grades). Researchers of doctor well-being were identified through academic conference programmes and abstracts, and authorship of relevant publications. Organisations measuring doctor well-being were identified through reports and publications on doctor well-being, discussions at national group meetings (i.e., Royal College of Physicians Flexibility and Wellbeing Group; BMA Wellbeing Support Stakeholder Group), and responses from a previous survey. ^27^ (A full list of organisations can be found in Supplementary Materials 2.) Professionals with experience managing doctor well-being were identified through policy documents and attendee lists from national well-being group meetings (i.e., Practitioner Health Programme; BMA Support). Doctors who had participated in the doctor involvement/engagement events or our online survey and had given permission to be approached were invited to participate. Additionally, and with the permission of the group administrator, doctors who were members of the BMA Wellbeing Stakeholder group, and Royal College of Physicians Flexibility and Wellbeing groups were invited to participate. We further identified stakeholders for all groups through recommendations from others. All potential participants were emailed an invitation to participate in the Delphi survey with a participant information sheet, a URL link to DelphiManager, and a link to the study information video. Participants were required to consent before registering their details (name and email). All participants were assigned a Study ID at registration, meaning data were anonymous at the point of collection.

### Delphi Survey and Analysis

The 43 well-being outcomes were listed with plain English descriptions by domain in a Delphi survey conducted over two rounds (Round 1 took place July 2021 and Round 2 August 2021). We asked participants to rate the importance of including each outcome in the COS using a 9-point Likert Scale. Ratings were grouped into three categories: a score of 1-3 on the Likert scale indicates the outcome is of ‘limited importance to include’, a score of 4-6 indicates the outcome is ‘important, but not critical’ for inclusion in the COS, and a score of 7-9 indicates that the outcome is critical to capture and measure in the COS. Participants had the option to declare they were unable to score an item. There was a free text comment box, and participants were encouraged to provide a rationale for their selection or additional comments. At the end of each Delphi Round, participants had the opportunity to suggest additional outcomes for possible inclusion in the COS; they were instructed that these should not be a symptom, sign or disease, nor a determinant of well-being. The criterion for considering an outcome for inclusion was that the definition of the outcome in the literature should differ conceptually from the help text offered for the existing outcomes. Where additional outcomes were suggested, the participant was emailed with the justification for inclusion or exclusion and given the opportunity to provide additional evidence/explanation.

In Round 2, the percentage of participants giving each rating for an outcome was fed back to participants. Participants were also reminded of their own ratings for each outcome from Round 1. Participants had the opportunity to re-rate each outcome based on this feedback. Participants were sent three reminder emails to complete Rounds.

The *a-priori* definition of consensus was when ≥75% of participants rated an outcome as ‘critical for inclusion’ (Ratings 7-9). This definition aligns with those used in other core outcome set development studies (e.g., ^46–49^) and the Grading of Recommendations Assessment, Development and Evaluation (GRADE) working group.^50^ The outcomes that met the threshold for inclusion in the COS were emailed to participants with an invitation to provide further comment on the core outcome set.

## RESULTS

### Phase 1. Generating a List of Outcomes

Our systematic review is described in detail elsewhere.^27^ A total of 78 verbatim well-being outcomes were extracted from the systematic review. To confirm uniqueness, these outcomes were grouped using definitions created from the research and policy literature [Supplementary Materials 1]. Thirty-four outcomes were found to meet the criteria for uniqueness. Of these 34 outcomes, 25 outcomes are synthesised; that is, they are comprised of multiple verbatim outcomes with the same conceptual definition (e.g., ‘health’, ‘self-esteem’). The remaining nine could not be grouped with any other verbatim outcomes and could only be described by a single term and definition (these included ‘Satisfaction with patient care’, ‘Psychological safety’ and ‘Compassion satisfaction’). No new outcomes were extracted from the doctor engagement/involvement events, and free text responses from the national survey, as these verbatim outcomes were all found to be among those already extracted from the systematic review. Next, the 34 unique outcomes were compared with existing frameworks and policy documents, which resulted in the identification of a further 9 outcomes, giving a final long list of 43 unique well-being outcomes (Table 1). This long list was then examined to identify whether sub-groups or themes existed between the outcomes. Five domains were identified: i) Overall appraisal of wellbeing, ii) Functional components of wellbeing, iii) Activity and participation components of wellbeing, iv) work-related wellbeing, and v) Health and social care specific wellbeing.

**Table 1.**
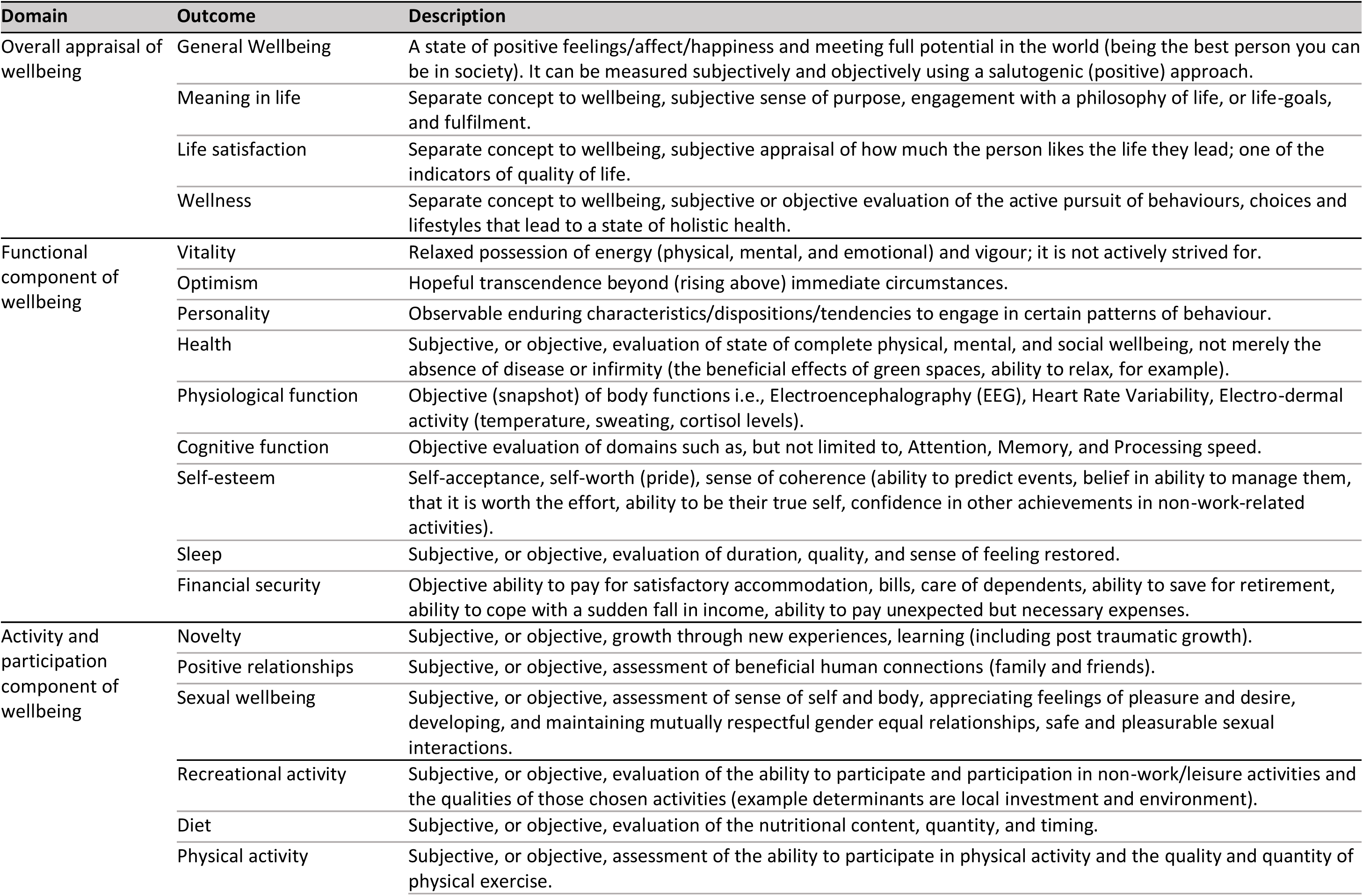
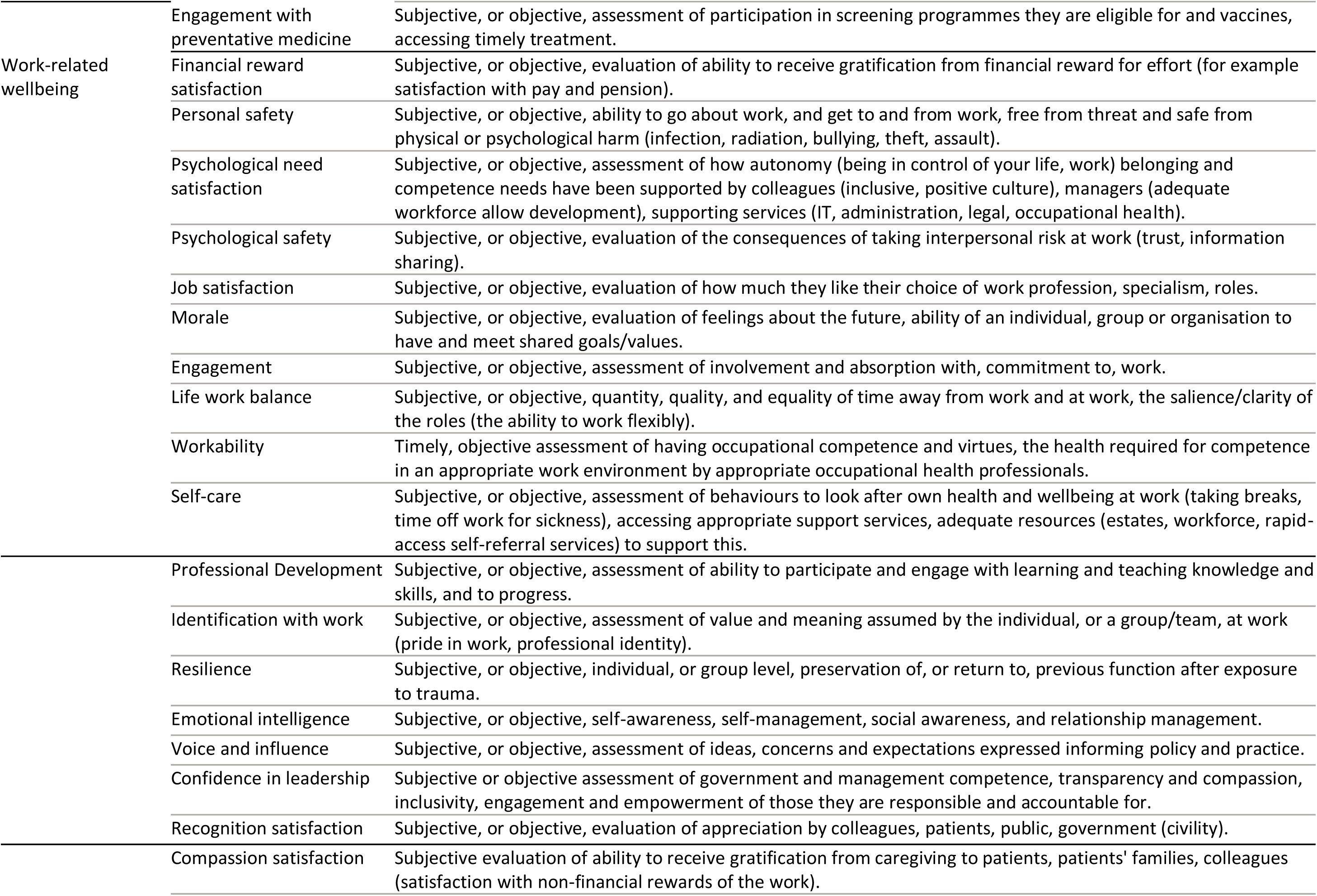
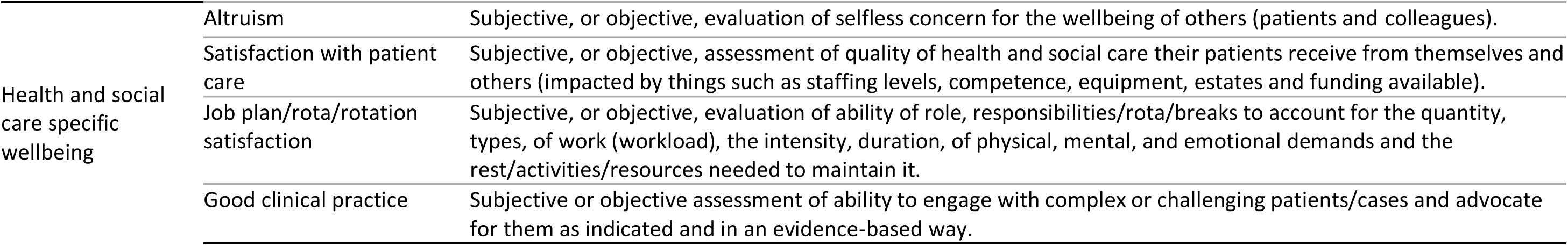
Outcomes and their descriptions by domain

### Phase 2. Delphi Survey

Invitations to participate were sent to 72 individuals across the four stakeholder groups. Sixty participants registered for the study: giving a response rate of 83.3%. One participant withdrew, meaning 59 participated in Round 1, of which 56 participants rated every outcome. Of the 59 participants, 52 also participated in Round 2: giving a retention rate from Round 1 to Round 2 of 91.2%. In Round 2, 51 participants rated every outcome. All rated outcomes were included in the analysis. Of these participants, 35% were male and 80% were doctors, of which 54% were consultants, 32% GPs, 7% Associates Specialists and 7% Training Grade Doctors.

In Round 1, 18 suggestions were made for additional outcomes from 8 participants (Supplementary Materials 3). None of these met the criteria for inclusion into Round 2. Of the suggested outcomes, 18 were not novel and had already been captured by existing outcomes, and 4 were interventions, 1 suggestion ‘Going on holiday’ was classified as both an intervention and not novel, being captured under ‘life work balance’ and ‘job plan/rota satisfaction’.

At the end of Round 2, seven well-being outcomes met the ≥75% threshold for inclusion in the Core Outcome Set for capturing and measuring doctor well-being: General well-being, Health, Personal Safety, Job satisfaction, Morale, Life work balance and Good clinical practice (Table 2). We sent the agreed set of outcomes to the stakeholder participants for review. They approved the COS without amendment.

**Table 2.** The final core outcome set.

| Domain | Outcome | Stakeholder rating outcomes as 'critical' % |
| --- | --- | --- |
| Overall appraisal | General wellbeing | 97.9 |
| Functional component of wellbeing | Health | 75.0 |
| Work-related wellbeing | Personal safety | 77.1 |
|  | Job satisfaction | 85.4 |
|  | Morale | 83.3 |
|  | Life work balance | 93.8 |
| Health and social care specific wellbeing | Good Clinical Practice | 89.6 |

## DISCUSSION

We have developed a core outcome set for capturing and measuring the well-being of doctors working in the NHS. By using a salutogenic and consensus approach, we have achieved agreement among four key stakeholder groups (Researchers, organisations that measure NHS doctor well-being, professionals with experience managing doctor well-being, and doctors). To our knowledge, this is the first time the COS-STAD guidance^28^ has been applied to the non-pathological concept of well-being, demonstrating that it is feasible to capture and measure. Our systematic review revealed heterogeneity between well-being outcomes and measurement tools.^27^ Implementing the COS will ensure a consistent and comprehensive focus for doctor well-being data-collection and prevent duplication, facilitate the generation of big data, and enable comparison at organisational, local and national levels. We recommend that future research, organisational and governance measurements of NHS doctors’ well-being use this COS. This does not preclude other outcomes from being measured as required but rather represents the minimum that should be captured and measured. Further research as to how best to operationalise and measure these well-being outcomes is now needed.

The long list of outcomes presented to stakeholders was evidence-based, drawing on doctor-specific research and the broader well-being literature.^27^ The finding that the listed outcomes already represented the additional well-being outcomes suggested by participants in Delphi Round 1 suggests that the list was inclusive and holistic. The seven outcomes of the COS can be mapped to existing doctor surveys. For example, the outcomes ‘General wellbeing’ and ‘Health’ are captured by the Office for National Statistics^51^ and the NHS staff survey;^52^ ‘Personal safety’ has been captured by the General Medical Council’s National Training Survey;^53^ ‘life work balance’ has been included in the NHS staff survey since 2021,^54^ mapping to the NHS People Plan promise theme of ‘We work flexibly.’ ^55^ Morale is also measured in the NHS staff survey.^52^ This suggests the COS is relevant and acceptable and that such data-capturing exercises would benefit from its implementation. It was noted that outcomes from the ‘Activity and Participation’ domain did not meet the threshold for inclusion. Outcomes under this domain include ‘positive relationships’, ‘recreational activity’, ‘diet’ and ‘physical activity’, also ‘engaging with preventative medicine’, including vaccination and medical screening. These outcomes require input and action from the individual, an understanding of the risk and severity of the threat to their well-being, and awareness of the benefits of acting.^56^ Outcomes included in the COS are arguably determined by the context or system in which an individual works. However, activity and participation outcomes were commonly studied in the literature as primary and secondary interventions, for example, mindfulness training and practices (e.g.,^57–61^) and debriefing ^62^ and dialogue groups.^63^ ^64^ Further consideration should be given to this domain in relation to doctor well-being. Good clinical practice has not been an outcome in existing surveys or data-collection exercises; this is a novel area of inquiry that requires further investigation.

There are a number of strengths to our study. To our knowledge, we developed the first COS for capturing and measuring doctor well-being and did so using the robust methodology set down in the COS-STAD guidelines.^28^ The COS is the product of consensus opinion from four stakeholder groups, including NHS doctors, but it is the involvement of representatives from professional bodies (i.e., British Medical Association, General Medical Council, Royal college of Physicians) that is a particular strength of this study. The uptake and use of a COS have traditionally been poor, and further research is needed to understand how uptake might be improved more broadly. ^65^ By including representatives from professional bodies, outcomes relevant to this group were considered in the creation of this COS. The number of participants in this study was acceptable for a Delphi study,^29^ and the >90% retention rate means attrition bias was not present. We ensured that the domains were not predetermined but led by the list of outcomes and their descriptions. However, the lack of an agreed definition of doctor well-being is a limitation. The published literature guided each outcome’s description and we found variation in how outcomes were described in the literature, with some being described by a single term and others by multiple terms. For example, the outcome ‘wellbeing’ had the most (n=19) non-unique terms extracted from the literature (i.e., General wellbeing, Personal wellbeing, Mental wellbeing, Physical wellbeing, Professional Wellbeing, Physician wellbeing, etc). Without an international consensus operational definition of doctor well-being, classification of outcomes and domains is subjective. To mitigate this, we published an operational definition of well-being^26^ derived from our literature review.^27^ Some outcomes posed a particular challenge as they did not fit the definition of well-being used for this study.^26^ Resilience is generally considered a ‘toxic term’ in medical culture.^66^ ^67^ However, it was included for completeness as it features so heavily in policy and research literature on doctor well-being.

This study aimed to create a COS for doctors working in the NHS, and accordingly, stakeholders were UK-based. While this COS might have some international relevance, it will be important to evaluate the acceptability and applicability of this COS for doctors in other countries, particularly where healthcare systems differ from the UK. The outcomes to emerge from the Delphi study, are not necessarily unique to doctors as research suggests that causes and interventions for poor mental well-being are not necessarily profession-specific.^68^ The robust methodology we have applied in this study could be repeated to establish the degree to which this COS could be used to capture and measure the well-being of other healthcare professionals. Our next step is to identify and agree on measurement tools and criteria. This would further enhance the quality and consistency of data collected using this COS.

## CONCLUSION

This COS is a stakeholder-derived set of outcomes that are most important to capture and measure for doctors’ well-being. Its application in research and health service governance will reduce heterogeneity and allow for better synthesis of evidence underpinning future workforce well-being policies and interventions, potentially improving doctor well-being and workforce retention. Research is needed to identify and evaluate outcome measurement tools.

**Twitter/X:** @C4WWellbeing

## Author Contributions

- GS designed the study and collected, analysed, and interpreted the data. Edited and approved the final article.
- NK wrote the manuscript, edited the article and approved the final article.
- DSB acquired funding, supervised the study’s design, data collection, analysis and interpretation, edited and approved the article and acted as guarantor.

## Disclaimer

The views expressed in this publication are those of the authors and not necessarily those of Health Education England, the National Institute for Health Research, or the Department of Health and Social Care.

## Data Availability

All data produced in the present work are contained in the manuscript and supplementary materials files.

## Acknowledgements

The authors would like to thank all those who participated in this Delphi study and supported the creation of this COS. Thanks to Dr Aimee O’Neill and Professor Julia Sinclair for their input into the early stages of the study and to Professor Peter Griffiths for reviewing the draft manuscript.

## Data availability statement

All data relevant to the study are included in the article or uploaded as supplementary materials.

