## Supplementary Material 1 for "Which outcomes should be included in a core outcome set for capturing and measuring doctor well-being? A Delphi study"

**Supplementary Materials 4**:

**Table A.** Outcome ratings from Delphi survey Round 1 (n = 59)

| Domain | Outcome | Limited Importance (%) | Important (%) | Critically Important (%) |
|---|---|---|---|---|
| Overall appraisal of wellbeing | General Wellbeing | 0.0 | 15.3 | 74.6 |
| | Meaning in life | 1.7 | 50.8 | 39.0 |
| | Life satisfaction | 3.4 | 42.4 | 45.8 |
| | Quality of life | 0.0 | 33.9 | 57.6 |
| | Wellness | 0.0 | 35.6 | 55.9 |
| Functional component of wellbeing | Vitality | 1.7 | 59.3 | 32.2 |
| | Optimism | 0.0 | 64.4 | 28.8 |
| | Personality | 20.3 | 57.6 | 13.6 |
| | Health | 0.0 | 32.2 | 62.7 |
| | Physiological function | 11.9 | 40.7 | 39.0 |
| | Cognitive function | 8.5 | 28.8 | 54.2 |
| | Self-esteem | 5.1 | 30.5 | 59.3 |
| | Sleep | 0.0 | 32.2 | 62.7 |
| | Financial security | 6.8 | 49.2 | 37.3 |
| Activity and participation component of wellbeing | Novelty | 23.7 | 59.3 | 11.9 |
| | Positive relationships | 6.8 | 32.2 | 61.0 |
| | Sexual wellbeing | 22.0 | 67.8 | 8.5 |
| | Recreational activity | 5.1 | 57.6 | 35.6 |
| | Diet | 5.1 | 66.1 | 28.8 |
| | Physical activity | 3.4 | 61.0 | 35.6 |
| | Engagement with preventative medicine | 20.3 | 54.2 | 23.7 |
| Work-related wellbeing | Financial reward satisfaction | 10.2 | 66.1 | 16.9 |
| | Personal safety | 3.4 | 22.0 | 67.8 |
| | Psychological need satisfaction | 0.0 | 33.9 | 57.6 |
| | Psychological safety | 5.1 | 33.9 | 55.9 |
| | Job satisfaction | 0.0 | 27.1 | 67.8 |
| | Morale | 0.0 | 28.8 | 66.1 |
| | Engagement | 0.0 | 54.2 | 40.7 |
| | Life work balance | 0.0 | 20.3 | 74.6 |
| | Workability | 5.1 | 52.5 | 23.7 |
| | Self-care | 0.0 | 39.0 | 52.5 |
| | Professional Development | 1.7 | 54.2 | 39.0 |
| | Identification with work | 10.2 | 61.0 | 20.3 |
| | Resilience | 5.1 | 52.5 | 33.9 |
| | Emotional intelligence | 6.8 | 52.5 | 33.9 |
| | Voice and influence | 8.5 | 47.5 | 37.3 |
| | Confidence in leadership | 1.7 | 47.5 | 45.8 |
| | Recognition satisfaction | 3.4 | 52.5 | 37.3 |
| Health and social care specific wellbeing | Compassion satisfaction | 1.7 | 45.8 | 44.1 |
| | Altruism | 11.9 | 62.7 | 20.3 |
| | Satisfaction with patient care | 0.0 | 42.4 | 54.2 |
| | Job plan/rota/rotation satisfaction | 0.0 | 40.7 | 55.9 |
| | Good clinical practice | 0.0 | 27.1 | 69.5 |

**Table B**. Outcome ratings from Delphi survey Round 2 (n = 52)

| Domain | Outcome | Limited Importance (%) | Important (%) | Critically Important (%) |
|---|---|---|---|---|
| Overall appraisal of wellbeing | General Wellbeing | 0.0 | 2.1 | 97.9 |
| | Meaning in life | 2.1 | 68.8 | 29.2 |
| | Life satisfaction | 2.1 | 60.4 | 37.5 |
| | Quality of life | 0.0 | 45.8 | 54.2 |
| | Wellness | 0.0 | 33.3 | 66.7 |
| Functional component of wellbeing | Vitality | 0.0 | 81.3 | 18.8 |
| | Optimism | 0.0 | 85.4 | 14.6 |
| | Personality | 12.5 | 83.3 | 4.2 |
| | Health | 0.0 | 25.0 | 75.0 |
| | Physiological function | 2.1 | 70.8 | 22.9 |
| | Cognitive function | 0.0 | 37.5 | 60.4 |
| | Self-esteem | 4.2 | 22.9 | 72.9 |
| | Sleep | 0.0 | 29.2 | 70.8 |
| | Financial security | 2.1 | 64.6 | 33.3 |
| Activity and participation component of wellbeing | Novelty | 16.3 | 79.6 | 4.1 |
| | Positive relationships | 0.0 | 34.7 | 65.3 |
| | Sexual wellbeing | 18.4 | 77.6 | 4.1 |
| | Recreational activity | 0.0 | 83.7 | 16.3 |
| | Diet | 0.0 | 83.7 | 16.3 |
| | Physical activity | 0.0 | 81.6 | 18.4 |
| | Engagement with preventative medicine | 12.2 | 77.6 | 10.2 |
| Work-related wellbeing | Financial reward satisfaction | 0.0 | 91.7 | 8.3 |
| | Personal safety | 0.0 | 22.9 | 77.1 |
| | Psychological need satisfaction | 0.0 | 31.3 | 66.7 |
| | Psychological safety | 2.1 | 50.0 | 47.9 |
| | Job satisfaction | 0.0 | 14.6 | 85.4 |
| | Morale | 0.0 | 16.7 | 83.3 |
| | Engagement | 0.0 | 64.6 | 35.4 |
| | Life work balance | 0.0 | 6.3 | 93.8 |
| | Workability | 2.1 | 75.0 | 16.7 |
| | Self-care | 0.0 | 47.9 | 50.0 |
| | Professional Development | 0.0 | 70.8 | 29.2 |
| | Identification with work | 2.1 | 83.3 | 14.6 |
| | Resilience | 4.2 | 62.5 | 33.3 |
| | Emotional intelligence | 2.1 | 66.7 | 31.3 |
| | Voice and influence | 0.0 | 60.4 | 39.6 |
| | Confidence in leadership | 0.0 | 31.3 | 68.8 |
| | Recognition satisfaction | 0.0 | 68.8 | 31.3 |
| | Compassion satisfaction | 2.1 | 68.8 | 29.2 |

| | | | | |
|---|---|---|---|---|
| Health and social care specific wellbeing | Altruism | 10.4 | 79.2 | 10.4 |
| | Satisfaction with patient care | 0.0 | 47.9 | 52.1 |
| | Job plan/rota/rotation satisfaction | 0.0 | 56.3 | 43.8 |
| | Good clinical practice | 0.0 | 10.4 | 89.6 |
