## Supplementary Material 2 for "Which outcomes should be included in a core outcome set for capturing and measuring doctor well-being? A Delphi study"

**Supplementary Material 3**: Additional outcomes suggested in Round 1 with reasons for not including in Round 2.

| | Suggested New Outcomes | Reasons for included/excluding |
|---|---|---|
| 1 | Psychological mentoring for every doctor as mandatory | **Intervention, not wellbeing outcome**. The ability to engage with this outcome and its effect would be captured by existing outcomes (Psychological safety, Psychological needs satisfaction). |
| 2 | Spirituality | **Not novel, captured by existing outcomes**. Spirituality was originally included as an outcome, but after piloting the Delphi it was suggested that more inclusive outcome of 'Optimism' to capture spiritual transcendence would be better and that religiosity would be captured in the existing outcome of 'Personality'. |
| 3 | Going on holiday | This could be **considered an intervention**, or if it is the ability to have time off work, it is **not novel, captured by existing outcomes** (Life work balance, Job plan/rota satisfaction). |
| 4 | Feel able to relax | **Not novel, captured by existing outcomes**. I.e., 'health', 'Psychological and cognitive function', but the example was added to the relevant help texts. |
| 5 | Family time | **Not novel, captured by existing outcomes**. Captured by the more general outcomes of 'life work balance', 'positive relationships' and 'job plan/rota satisfaction'. |
| 6 | Sense of pride | **Not novel, captured by existing outcomes**. Captured by 'self-esteem', pride was added to the relevant help text. |
| 7 | Success, of confidence in other achievements in non-work-related activities | **Not novel, captured by existing outcomes**. Captured by 'self-esteem', example as added to the relevant help texts. |
| 8 | Post-traumatic growth | **Not novel, captured by existing outcomes.** To keep the number of outcomes as brief as possible, the concepts were as inclusive as possible. This suggestion was added to the help text for the outcome 'Novelty' to read subjective growth through new experiences, learning (including post-traumatic growth). |
| 9 | Sense of belonging | **Not novel, captured by existing outcomes.** Captured by 'Psychological need satisfaction' with the help text: *Subjective or objective assessment of how autonomy, belonging and competence needs have been supported by colleagues*. |
| 10 | Empowered to speak up (no microaggression) | **Not novel, captured by existing outcomes.** Captured by the outcomes 'Psychological Needs Satisfaction and 'Voice and influence'. |
| 11 | Flexibility of workplace | **Not novel, captured by existing outcomes.** Mentioned in the help text for 'life work balance' which reads "*subjective or objective quantity, quality and equality of time away from work and at work, the salience/clarity of roles (the ability to work flexibly)"* |

| | Suggested New Outcomes | Reasons for included/excluding |
|---|---|---|
| 12 | Compulsory day out and team building | **It is an intervention, not a wellbeing outcome**. The outcomes listed would captured the effect of these activities. |
| 13 | Family cared for so you can go to work and concentrate | **It is an intervention and not novel, captured by existing outcomes.** The outcomes 'Life work balance', 'confidence in leadership' and 'Financial security' would cover the time, infrastructure and finances that would be required to achieve this. |
| 14 | Security – as a wellbeing outcome when Maslow's basic human needs – warmth, food, shelter, plus security of meaningful employment – are met. I did not think any of the other categories quite captured the inability to feel wellbeing if you home or job is under threat. | **Not novel, Captured by existing outcomes.** This is captured by the outcome 'Financial security' which has the help text "*objective ability to pay for satisfactory accommodation, bills, care of dependents, ability to save for retirement, ability to cope with sudden fall in income, ability to pay unexpected, but necessary expenses".* |
| 15 | Political security – feeling that the NHS us valued and appropriately funded/resourced with well-paid colleagues. | **Not novel, captured by existing outcomes.** i.e., 'Voice and influence', 'Confidence in leadership', 'Satisfaction with patient care', 'Financial reward satisfaction'. The word Government was added to the help text for 'Confidence in leadership' to better cover this point, and to read "*Subjective or objective assessment of government and management competences, transparency and compassion, inclusivity, engagement and empowerment of those they are responsible and accountable for*." |
| 16 | Pride in work | **Not novel, captured by existing outcomes.** The help text for 'self-esteem' was altered to include: 'Sense of pride outside of work, as well as at work." |
| 17 | Decision making about career development | **Not novel, captured by existing outcomes.** Covered by the outcome 'Professional development', the help text of which reads "subjective or objective assessment of ability to participate and engage with learning and teaching knowledge and skills, and to progress". |
| 18 | Feeling in control of your life | **Not novel, captured by exiting outcomes.** The suggestion 'Feeling in control of your life' was added to the help text for 'Psychological needs satisfaction' to include "being in control of your life, work." |
