## Supplementary Material 3 for "Which outcomes should be included in a core outcome set for capturing and measuring doctor well-being? A Delphi study"

**Supplementary Materials 2**

Stakeholder organisations

- The symbol ≥ is used for n as the classification of the individuals and organisations into stakeholder groups was undertaken by the author.

1. Those who could use COS-DR in their research - based on that, they have previously published doctor well-being research individually (n≥15) or as an organisation (n≥5).

- British Medical Association
- Health Education and Improvement Wales
- Health Education England
- Kings' Fund
- Practitioner Health Programme

2. Organisations that measure doctor well-being in the NHS in the UK every year (n≥3).

- British Medical Association
- General Medical Council
- Practitioner Health Programme
- Royal College of Physicians

3. Professionals with experience of managing doctor wellbeing individually, including doctors, nurses, psychologists, and liaison advisors (n≥8) and organisations (n≥2).

- British Medical Association
- Practitioner Health Programme

4. Doctors individually (n=48) and organisations representing them (n≥12)

- Association of Anaesthetists
- British Association of Physicians of Indian Origin
- British Medical Association
- Faculty of Occupational Medicine
- Health Education and Improvement Wales
- Health Education England
- Kings' Fund
- Practitioner Health Programme
- Royal College of General Practitioners
- Royal College of Paediatric and Child Health
- Royal College of Physicians.
