## Supplementary Material 4 for "Which outcomes should be included in a core outcome set for capturing and measuring doctor well-being? A Delphi study"

### Supplementary Materials 1.

**Table A**. Outcomes identified through synthesis with the reasons they are not unique concepts (n=25).

| Unique Concept | Definition | Verbatim Outcomes from the Systematic Review | Reason not unique |
|---|---|---|---|
| Wellbeing | A state of positive feelings and meeting full potential in the world (being the best person you can be in society). It can be measured subjectively and objectively using a salutogenic (positive) approach. [1] | Overall wellbeing | Context adjective |
| | | General wellbeing | Context adjective |
| | | Self-perceived | Methodological adjective (how) |
| | | Subjective wellbeing | Methodological adjective (how) |
| | | Personal wellbeing | Context adjective |
| | | Individual wellbeing | Context adjective |
| | | Happiness | Hedonic component |
| | | Mental wellbeing | Artificial subgroup |
| | | Emotional wellbeing | Artificial subgroup |
| | | Psychological wellbeing | Artificial subgroup |
| | | Physical wellbeing | Artificial subgroup |
| | | Occupational wellbeing | Context adjective |
| | | Professional wellbeing | Context adjective |
| | | Job related wellbeing | Context adjective |
| | | Work related wellbeing | Context adjective |
| | | Physician wellbeing | Methodological adjective (who) |
| | | Resident wellbeing | Methodological adjective (who) |
| | | Organisational wellbeing | Methodological adjective (who) |
| | | Unit wellbeing | Methodological adjective (who) |
| Life satisfaction | Separate concept to wellbeing, subjective appraisal of how much the person like the life they lead; one of the indicators of quality of life. [2 3] | Satisfied with life | Phrasing |
| Quality of life | Separate concept to wellbeing, subjective appraisal of individuals positioned in life in the context of the culture and value systems in which they live and in relation to their goals, expectations, standards, and concerns. [4-6] | Social quality of life | Artificial subgroup |
| | | Environmental quality of life | Artificial subgroup |
| Wellness | | Health behaviours | Component |

| Unique Concept | Definition | Verbatim Outcomes from the Systematic Review | Reason not unique |
|---|---|---|---|
| | Separate concept to wellbeing, subjective or objective evaluation of active pursuits of behaviours, choices and lifestyles that lead to a state of holistic health. [7] | Physician wellness | Methodological adjective |
| Optimism | Hopeful transcendence beyond (rising above) immediate circumstances.[8] | Spiritual wellbeing | Subgroup of wellbeing Definition of spiritualism the same as optimism, if religion is left out [559] |
| Health | Subjective or objective evaluation of state of complete physical, mental and social wellbeing, not merely the absence of disease of infirmity (the beneficial effects of green spaces for example). [9, 10] | Mental health | Artificial subgroup |
| | | Psychological health | Artificial subgroup |
| | | Self-perceived health | Methodological adjective |
| | | Satisfaction with health | Methodological adjective |
| | | General health status | Phrasing |
| Cognitive function | Objective evaluation of domains such as, but not limited to, Attention, Memory, Processing speed. [11, 12] | Mental energy | Methodology-self report measure |
| Self-esteem | Self-acceptance, self-worth, sense of coherence (ability to predict events, belief in ability to manage them, that it is worth the effort, ability to be their true self). [13] | Confidence | Component |
| | | Professional self esteem | Context adjective |
| | | Global self esteem | Context adjective |
| Sleep | Subjective or objective evaluation of duration, quality, and sense of feeling rested. [14] | Sleep problems | Methodology - pathogenic |
| Financial security | Objective ability to pay for satisfactory accommodation, bills, care of dependents, ability to save for retirement, ability to cope with a sudden fall in income, ability to pay unexpected, but necessary expenses. [15] | Household economic wellbeing | Wellbeing subgroup |
| Positive relationships | Subjective or objective assessment of beneficial human connections. [16] | Satisfaction with family relations | Methodology |
| | | Satisfaction with family support | Methodology |
| Psychological need satisfaction | Subjective or objective assessment of how autonomy, belonging and competence needs have been supported by colleagues (inclusive, positive culture), managers (adequate workforce to allow development), supporting services (IT, administration, legal, occupational health). [17] | Job demand support control model | Component |
| Job satisfaction | | Occupational choice satisfaction | Methodology |

| Unique Concept | Definition | Verbatim Outcomes from the Systematic Review | Reason not unique |
|---|---|---|---|
| | Subjective or objective evaluation of how much they like their choice of work profession, specialism, roles. [18] | Work satisfaction | Synonym |
| | | General job satisfaction | Context adjective |
| | | Career satisfaction | Synonym |
| | | Professional satisfaction | Synonym |
| Morale | Subjective or objective evaluation of feelings about the future ability of an individual, group or organisation to have and meet shared goals/values. [19] | Career morale | Context adjective |
| | | Speciality morale | Context adjective |
| | | Participation | Methodology |
| | | Goal clarity | Component |
| Engagement | Subjective or objective assessment of involvement and absorption with, commitment to, work. [20] | Motivational effort | Methodology |
| | | Employee job intentions | Methodology |
| Altruism | Subjective or objective evaluation of selfless concern for the wellbeing of others. [21] | Altruistic behaviour | Methodology |
| Life work balance | Subjective or objective quantity, quality, and equity of time away from work and at work, the salience/clarity of the roles (the ability to work flexibly). [22-24] | Satisfaction with work-life balance | Methodology |
| | | Work rest balance | Phrasing |
| | | Work pressure | Component |
| | | Role behaviour | Methodology |
| | | Salience with life roles | Component |
| Professional Development | Subjective or objective assessment of ability to participate and engage with learning and teaching knowledge and skills, and to progress. [25] | Skills development | Context adjective |
| | | Efficiency | Methodology |
| Identification with work | Subjective or objective assessment of value and meaning assumed by the individual, or a group, at work. [18] | Psychological identification with work | Artificial subgroup |
| | | Meaning at work | Component |
| | | Meaning in work | Component |
| Resilience | Subjective or objective individual, or group level, preservation of, or return to, previous function after exposure to trauma.[26] | Strain resistant resources | Components |
| Voice and influence | Subjective or objective assessment of ideas, concerns and expectations expressed informing policy and practice.[27] | Influence | Component |
| Confidence in leadership | Subjective or objective assessment of management competence, transparency and compassion, inclusivity, | Supportive leadership | Component |
| | | Authority | Synonym |

| Unique Concept | Definition | Verbatim Outcomes from the Systematic Review | Reason not unique |
|---|---|---|---|
| | engagement and empowerment of those they are responsible and accountable for. [27] | Perceived organisational support | Context component |
| Recognition satisfaction | Subjective or objective evaluation of appreciation by colleagues, patients, public, government (civility, ward rounds/appointments attended and on time). [28 29] | Feedback | Methodology |
| Job plan/rota/rotation satisfaction | Subjective or objective evaluation of ability of job plan/rota/rotation to account for the quantity, types, of work (workload), the intensity, duration, of physical, mental and emotional demands and time rest/activities/resources needed to maintain it. [30] | Workload | Component |
| Good clinical practice | Subjective or objective assessment of ability to engage with high-risk cases, follow standards where appropriate not rigidly, use diagnostic tests and treatment when clinically indicated and evidence based, not just in case. [31] | Self-reported competence | Methodology |

**Table B**. Unique outcomes for which there were no other terms (n=9)

| Unique concept | Definition |
|---|---|
| Sexual wellbeing | Subjective, or objective, assessment of sense of self and body, appreciating feelings of pleasure and desire, developing and maintaining mutually respectful gender equal relationships, safe and pleasurable sexual interactions.[32] |
| Personality | Observable enduring characteristics/dispositions/tendencies to engage in certain patterns of behaviour.[8] |
| Diet | Subjective, or objective, evaluation of the nutritional content, quantity, and timing.[33] |
| Physical activity | Subjective, or objective, assessment of the ability to participate in physical activity and the quality and quantity of physical exercise. [34] |
| Physiological functioning | Objective (snapshot) of body function i.e., Electroencephalography (EEG), Heart Rate Variability, Electro-dermal activity (temperature, sweating), hypothalamic-pituitary axis hormones. [11 35 36] |
| Workability | Timely, objective assessment of having occupational competence and virtues, the health required for competence in an appropriate work environment by appropriate occupational health professionals.[37] |
| Compassion Satisfaction | Subjective evaluation of ability to receive gratification from caregiving to patients, patients' families, colleagues (satisfaction with non-financial rewards of the work).[38] |

| | |
|---|---|
| Psychological safety | Subjective or objective evaluation of the consequences of taking an interpersonal risk at work (trust, information sharing).[39] |
| Satisfaction with patient care | Subjective, or objective assessment of quality of health and social care their patients receive. |

**Table C.** List of unique outcomes identified from other sources (n=9)

| Unique concept | Definition | Sources |
|---|---|---|
| Meaning in life | Separate concept to wellbeing, subjective sense of purpose, engagement with a philosophy of life or life-goals, fulfilment.[40 41] | WHOQOL SRPB[42]<br>European Social Survey wellbeing module[43] |
| Vitality | Relaxed possession of energy (physical, mental and emotional) and vigour, it is not actively strived for.[44] | European Social Survey wellbeing module[43]<br>Wilson and Cleary framework (1995)[5 45]<br>WHO ICF[46] |
| Novelty | Subjective or objective, growth through new experiences, learning.[47] | European Social Survey wellbeing module[43]<br>[from Doctor Engagement Event] |
| Recreational activity | Subjective, or objective, evaluation of the ability to participate and participation in non-work/leisure activities and the qualities of those chosen activities.[48] | Diener et al (1999) [45] |
| Engagement with preventative medicine | Subjective, or objective, assessment of participation in screening programmes they are eligible for and vaccine, timely treatment. | Outcomes for Graduates[49]<br>NHS People Plan[27] |
| Financial reward satisfaction | Subjective, or objective, evaluation of ability to receive gratification from financial reward for effort. [50 51] | Contract and pension disputes[52 53] |
| Personal safety | Subjective, or objective, ability to go about work, and get to and from work, free from threat and safe from physical or psychological harm (infection, radiation, bullying, harassment, theft, assault). | NHS People Plan[27] |
| Self-Care | Subjective, or objective, assessment of behaviours to look after own health and wellbeing at work (taking breaks, time off work for sickness), accessing appropriate support services, adequate resources (estates, workforce, rapid-access self-referral services) to support this. | Outcomes for Graduates[49] |
| Emotional Intelligence | Subjective, or objective, self-awareness, self-management, social-awareness, and relationship management.[54 55] | Building and strengthening leadership[56] |
